# Multi-level Predictors of Depression Symptoms in the Adolescent Brain Cognitive Development (ABCD) Study

**DOI:** 10.1101/2021.02.11.21251432

**Authors:** Tiffany C. Ho, Rutvik Shah, Jyoti Mishra, April C. May, Susan F. Tapert

## Abstract

**Objective:** To identify multi-level factors that maximize prediction of depression symptoms in a diverse sample of children in the U.S. participating in the Adolescent Brain and Cognitive Development (ABCD) study.

**Methods:** 8,507 participants (49.6% female, 75.2% white, ages 9-10) from ABCD provided complete data at baseline and 7,998 of these participants provided one-year follow-up data. Depression symptoms were measured with the Child Behavior Checklist. Predictive features included child demographic, environmental, and structural and resting-state fMRI variables, parental depression symptoms and demographic characteristics, and relevant site and scanner-related covariates. We used linear (elastic net regression, EN) and non-linear (gradient boosted trees, GBT) predictive models to identify which set of features maximized prediction of depression symptoms at baseline and, separately, at one-year follow-up.

**Results:** Both linear and non-linear models achieved comparable results for predicting baseline (EN: MAE=3.628; *R*^*2*^=0.232; GBT: MAE=3.555; *R*^*2*^=0.229) and one-year follow-up (EN: MAE=4.116; *R*^*2*^=0.143; GBT: MAE=4.141; *R*^*2*^=0.1400) depression. Parental depression symptoms, family support, and child sleep duration were among the top predictors of concurrent and future child depression symptoms across both models. Although resting-state fMRI features were relatively weaker predictors, connectivity of the right caudate was consistently the strongest neural feature associated with depression symptoms at both timepoints. In contrast, brain features derived from structural MRI did not significantly predict child depression symptoms.

**Conclusions & Relevance:** Parental mental health and child sleep quality are potentially modifiable risk factors for youth depression. Functional connectivity of the caudate is a relatively weaker predictor of depression symptoms but may represent a biomarker of depression risk.

## Introduction

Depression is a leading contributor of disability worldwide, increasingly prevalent, and highly recurrent (1). Its onset typically occurs during adolescence (2, 3); moreover, because adolescent-onset depression is associated with more severe functional impairment, greater risk for other mental and physical illnesses, and more recurrent episodes that are often resistant to treatment (4, 5), identifying risk factors for adolescent depression is critical for early intervention. While research studies have identified multiple risk factors for depression in adolescence that span different domains, including demographic, clinical, psychosocial, and neurobiological factors (6–11), most studies have sought to understand the role of isolated factors that each explain a small portion of variance rather than maximizing prediction across a broad set of factors.

Two analytic barriers have hindered our ability to integrate across a diverse set of distinct yet related risk factors: 1) large sample sizes are needed to accommodate statistical models with more predictors; and 2) risk factors for depression are often highly collinear. In this context, Adolescent Brain Cognitive Development (ABCD), an ongoing multi-site longitudinal study of brain development and mental health in nearly 12,000 U.S. children ages 9-10 collecting comprehensive demographic, clinical, psychosocial, and neurobiological information, represents an ideal opportunity to leverage machine learning methods that address instances of multicollinearity. Here, we utilized two complementary machine learning techniques—regularized linear regressions (elastic-net) and non-linear ensemble learning (gradient boosted trees) to identify the strongest statistical predictors of depression symptoms in the first waves of ABCD.

Because prior work from “big data” consortia have focused on task-independent MRI markers of depression, including morphological characteristics (i.e., surface area, cortical thickness) derived from structural MRI (12–15) and intrinsic resting-state functional connectivity (FC) obtained from fMRI signals (16, 17), we limited our feature set (i.e., statistical predictors) of brain-based variables to those derived from these imaging modalities for the purposes of comparability with other large-scale efforts, as well as for computational tractability and potential clinical utility.

Based on previous studies examining factors associated with the onset and presence of depression in adolescence (3, 18–21), we hypothesized that female gender, parental depression, morphology and functional connectivity of the amygdala, hippocampus, nucleus accumbens, as well as cortical regions involved in emotion regulation and self-referential processing will constitute the most important features in the best performing models. We also explored whether the top-performing neural features predicted depression symptoms in a subset of participants with follow-up data on depression symptoms one year later.

## Methods

### Participants

Baseline and follow-up data used for the present investigation were obtained from the Annual Curated Data Release 3.0 from the ABCD consortium (https://abcdstudy.org/index.html), which is a population-based cohort of 11,878 children ages 9–10 years recruited from 21 sites throughout the U.S. (22). Each recruitment site obtained full assent and consent from the children and their parent(s)/legal guardian(s), respectively in accordance with local Institutional Reviews Boards. Out of the 11,878 children with data at baseline, we excluded those without usable structural and functional MRI based on initial quality control screening, with familial participants (e.g., twins), and those with substantial missing postprocessed data, resulting in 8,507 unique participants at baseline. Of these, 7,998 provided one-year follow-up Child Behavior Checklist (CBCL) data. Please see **Figure S1** for a flowchart showing initial sample size and final sample size.

### Depression Symptoms (Outcome Measure)

Children and their parent/guardian completed the computerized Kiddie-Schedule for Affective Disorders–5 (K-SADS-5; 23–25) to assess current and lifetime history of Axis I disorders. Based on endorsement from either child- or parent-report, only 6.3% of the sample met lifetime criteria for any depressive disorder (i.e., Major Depressive Disorder, dysthymia, and depression not otherwise specified). Because history of depression does not necessarily reflect current depressive symptomatology and because classification models for imbalanced datasets tend to introduce bias and misclassification of the minority (less represented) class (26), we elected to assess current depression symptoms dimensionally. Specifically, we used scores from the “Depressive Problems” subscale of the Child Behavior Checklist (CBCL), which comprises items consistent with the DSM-5 criteria for depression in youth from parent report. The CBCL is one of the most widely used measures of emotional problems and provides standardized scores based on national norms in children ages 6-18 (27).

### Features (Predictors)

Non-brain features included parental depression symptoms, as measured by the Adult Self-Report (28), parent-reported demographic information (including child age, child sex assigned at birth, child race, child education level, parental marital status, parental income, and parental education), average hours of sleep per night (herein referred to as sleep duration), substances taken in the past 24 hours, site, scanner type (Siemens, GE, or Philips), and MRI device serial number.

All structural MRI data were preprocessed using FreeSurfer v. 5.30 (29) (http://surfer.nmr.mgh.harvard.edu/) and underwent automated and manual quality control procedures. All resting-state fMRI data were preprocessed using AFNI (30) and rigorous motion censoring was applied. See (31) and “Image Processing” in the Supplement for more details.

Brain features derived from structural MRI included subcortical gray matter volumes, cortical thickness, and cortical surface area from regions defined by the Desikan atlas (32). Because over 100 subjects were missing data from morphometry metrics based on the ventricles, we did not include ventricles in our analyses (see “Missing Data” in the Supplement). Participants with low quality control scores on their MRIs were excluded from all analyses (see “Image Processing” in the Supplement).

Brain features derived from resting-state fMRI included functional connectivity between networks defined by the Gordon parcellation (33) and also between each of the subcortical structures segmented by FreeSurfer with each of the cortical networks (see 28 for more details). These cortical networks included the auditory network, the cingulo-parietal network, the cingulo-opercular network, the default mode network, the frontoparietal network, the retrosplenial-temporal network, the ventral attention network, and the visual network. Participants for whom more than 10% of their timeseries framewise displacement > 0.2 mm or for whom there was fewer than 375 usable timepoints for modeling were excluded from the present analyses (see “Image Processing” in the Supplement for more details). See **Table S1** for a summary of the filenames and variables used as features and outcomes in our models and “Features (Predictors)” in the Supplement for more details.

### Machine Learning Analyses

All features were standardized prior to analysis using a robust scaler method (i.e., centered on the median and scaled by interquartile range) to resist influence of outliers. We used elastic net (EN) to perform regularized linear regression, which combines L1 (least absolute shrinkage and selection operator, or LASSO), and L2 (ridge) penalties (34) and histogram gradient boosted trees (light GBT), an ensemble method of supervised machine learning that does not assume linearity (35), using the *sklearn* package in Python. We first randomly split the data into 4-folds (25% data in each fold). Three randomly chosen folds (75% of the total data) was used for training (i.e., hyperparameter tuning and validation). The remaining 25% of the data was used solely for testing model performance, thereby maintaining independence in our training and test sets. In the training data (75% of the total data), we conducted hyperparameter tuning, training and validation over a 10-fold cross-validation scheme. In other words, 9-folds (67.5% of the total data) were used to identify a set of hyperparameters, using grid search, that best fit each model based on minimizing mean absolute error (MAE), and 1-fold (7.5% of the total data) was used for validation. This process was repeated ten times for each fold to identify a model with the optimal set of hyperparameters and then applied to the training set (75% of the total data). This final model was then evaluated on the remaining left-out test set (25% of the total data), from which we report our model performance metrics. See **Figure 1**.

**Figure 1.**
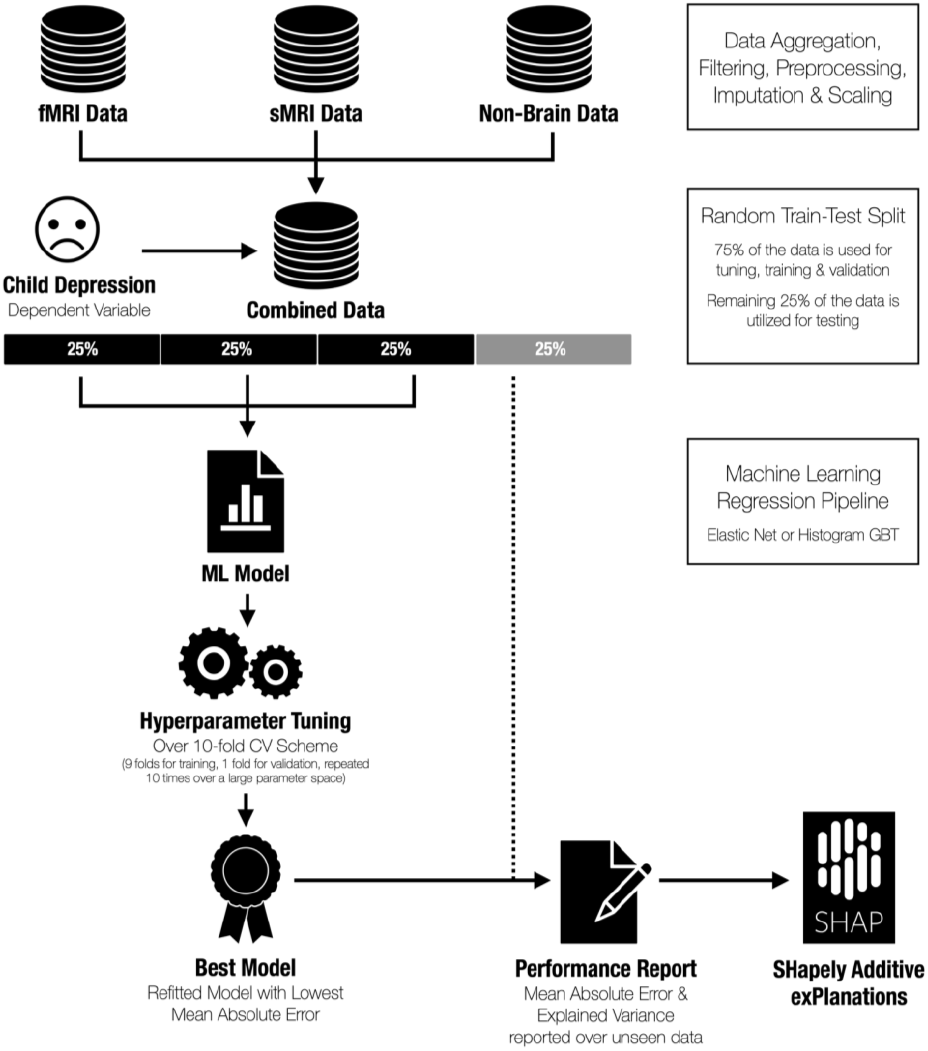
**Visualization of the machine learning approach used to identify features contributing to the prediction of concurrent depression symptoms.**

To determine the importance of each relative feature within the best performing model, we used the Shapley Additive exPlanations (SHAP) method (36). Briefly, a Shapley value for a feature quantifies how well a particular feature contributes to overall model performance even in the presence of correlated features. SHAP is a computationally efficient way to generate Shapley values, given the number of features in our models (almost 400 total). See “Feature Importance” in the Supplement for more details.

### Code Availability

All code for performing data cleaning, organization, and predictive modeling can be found here: https://github.com/tiffanycheingho/ABCD

## Results

The final analytic sample contained 8,507 participants: 49.6% were assigned female at birth and 75.2% identified as being White. Distributions of parental income and educational attainment were consistent with percentages reported from recent epidemiological data (37, 38). Please see **Table 1** for a summary of the descriptive statistics of the final sample at baseline. Please see **Table S1** for a summary of the descriptive statistics in the one-year follow-up (which were comparable to the full baseline sample).

**Table 1.** Demographics of final analytic sample (N=8,507). Continuous variables are reported as mean ± standard deviation (min – max). All variables reported here are at baseline.

|  | **Full sample** |
| --- | --- |
| **N** | 8,507 |
| **Sex (% Female)** | 49.65% |
| **Age (years)** | 9.49 ± 0.51 (8 – 11) |
| **Race (% White)** | 75.23% |
| **Puberty Development Score** | 6.77 ± 2.89 (1 – 20) |
| **Body mass index** | 18.98 ± 32.75 (2.08 - 52.82) |
| **% with caffeine consumption in past 24 hours** | 17.85% |
| **% with prescription medication use in past 24 hours** | 14.04% |
| **% with over-the-counter medications use in past 24 hours** | 14.85% |
| **Sleep duration (hours per night)** | 9-11 hours: 4072 (47.87%)<br>8-9 hours: 3181 (37.39%)<br>7-8 hours: 969 (11.39%)<br>5-7 hours: 263 (3.09%)<br><5 hours: 22 (0.26%) |
| **Current grade** | 1<sup>st</sup> grade: 0.025%<br>2<sup>nd</sup> grade: 0.45%<br>3<sup>rd</sup> grade: 17.03%<br>4<sup>th</sup> grade: 44.62%<br>5<sup>th</sup> grade: 34.71%<br>6<sup>th</sup> grade: 3.14%<br>7<sup>th</sup> grade: 0.025% |
| **Parental marital status** | Married: 69.39%<br>Never Married: 11.66%<br>Divorced: 8.73%<br>Living with Partner: 5.65%<br>Separated: 3.79%<br>Widowed: 0.78%<br>[60] |
| **Combined family income, past year** | <\$5k: 3.08%<br>\$5k - \$12k: 3.25%<br>\$12k - \$16k: 2.45%<br>\$16k - \$25k: 4.28%<br>\$25k - \$35k: 5.70%<br>\$35k - \$50k: 7.81%<br>\$50k - \$75k: 12.61%<br>\$75k - \$100k: 22%<br>\$100k - \$200k: 28.06%<br>>\$200k: 10.76% |
| **Highest parental education** | 3<sup>rd</sup> grade: 0.03%<br>4<sup>th</sup> grade: 0.02% |
|  | 5<sup>th</sup> grade: 0.02%<br>6<sup>th</sup> grade: 0.38%<br>7<sup>th</sup> grade: 0.11%<br>8<sup>th</sup> grade: 0.34%<br>9<sup>th</sup> grade: 0.73%<br>10<sup>th</sup> grade: 0.71%<br>11<sup>th</sup> grade: 0.99%<br>12<sup>th</sup> grade (no diploma): 1.39%<br>High school graduate: 6.62%<br>GED or equivalent: 2.62%<br>Some College Degree: 12.80% Associate Degree (Occupational, Technical, or Vocational): 7.41%<br>Associate Degree (Academic Program): 5.2%<br>Bachelor's Degree: 25.30%<br>Master's Degree: 24.40% Professional School Degree: 5.17%<br>Doctoral Degree: 5.76% |
| **Weekend daily screen time (hours)** | 3.80 ± 2.61 (0 – 24) |
| **CBCL (Youth) Depression (t-score)** | 53.56 ± 5.69 (50 – 89) |
| **ASR (Parent) Depression (t-score)** | 53.43 ± 5.70 (50 – 98) |

Both EN and GBT models yielded comparable results in terms of model performance (see **Table 2**). Across both models, parental self-report symptoms of depression at baseline was the strongest predictor of youth depression symptoms at baseline and one-year follow-up. Both models also identified prescription medicate usage, sleep duration, and greater family conflict as important features, although the rank order of these features in their contribution to their respective models differed slightly (see **Figures 2-3**).

**Table 2.** Summary of model performance metrics. EN=Elastic Net; GBT=Gradient Boosted Trees; MAE=Mean Absolute Error.

| **Outcome** | **Model Type** | **MAE** | **R<sup>2</sup>** |
| --- | --- | --- | --- |
| Baseline CBCL | EN (linear) | **3.57** | **0.23** |
| Baseline CBCL | GBT (non-linear) | **3.55** | **0.23** |
| Follow-up CBCL | EN (linear) | **4.12** | **0.16** |
| Follow-up CBCL | GBT (non-linear) | **4.14** | **0.14** |

**Figure 2.**
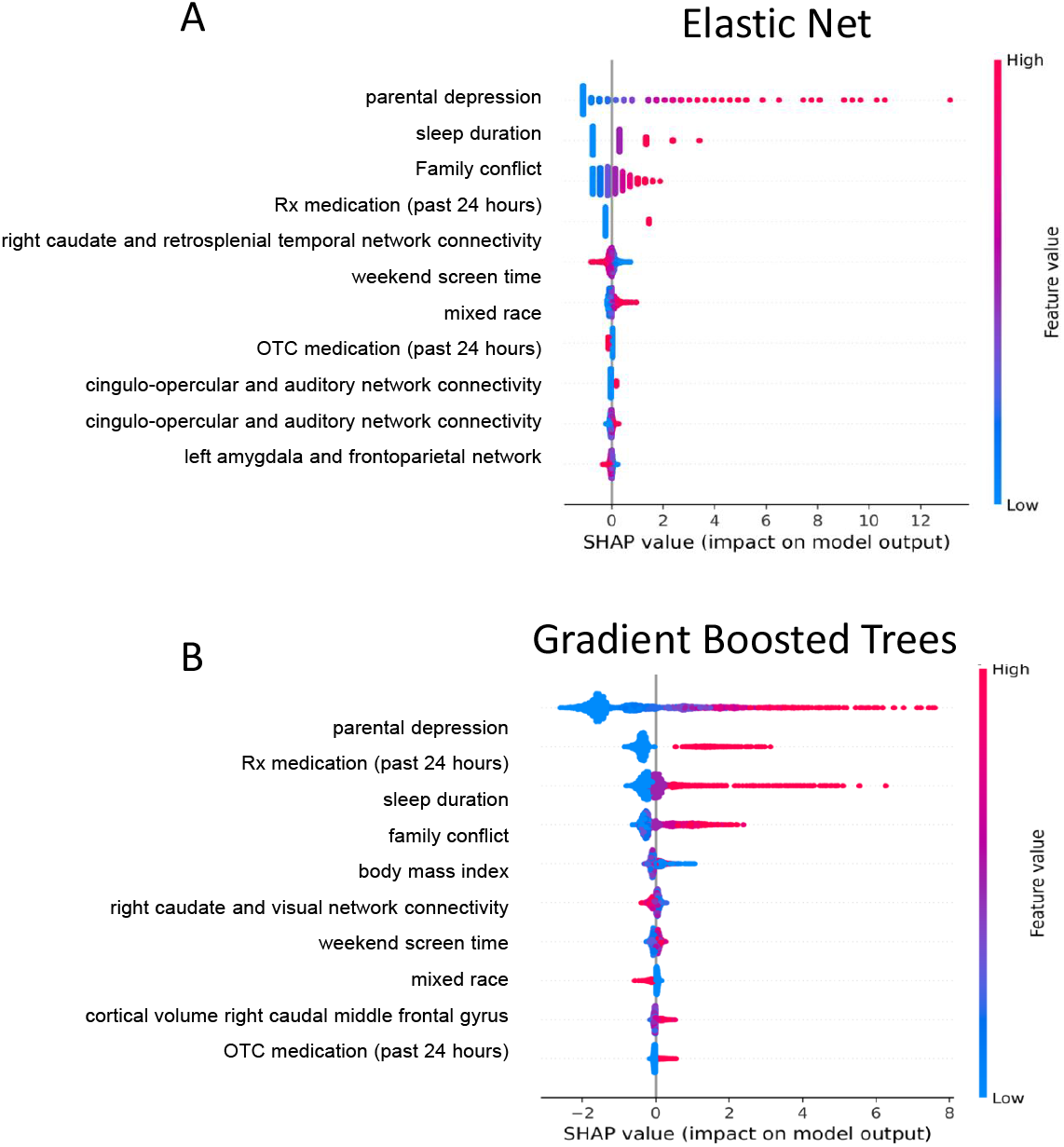
Shapley values of the top 10 features in the elastic net (A) and gradient boosted trees (B) models predicting baseline depression symptoms. The summary plots indicate the relationship between the value of a feature and the impact on the prediction, thus combining feature importance with feature effects. Each point on the summary plot is a Shapley value for a feature and an instance. The position on the y-axis is determined by the feature and on the x-axis by the Shapley value. The color represents the value of the feature from low (blue) to high (pink). The features are ordered according to their importance. See **Figure S3** for a summary of the magnitude of Shapley values per feature in each model.

**Figure 3.**
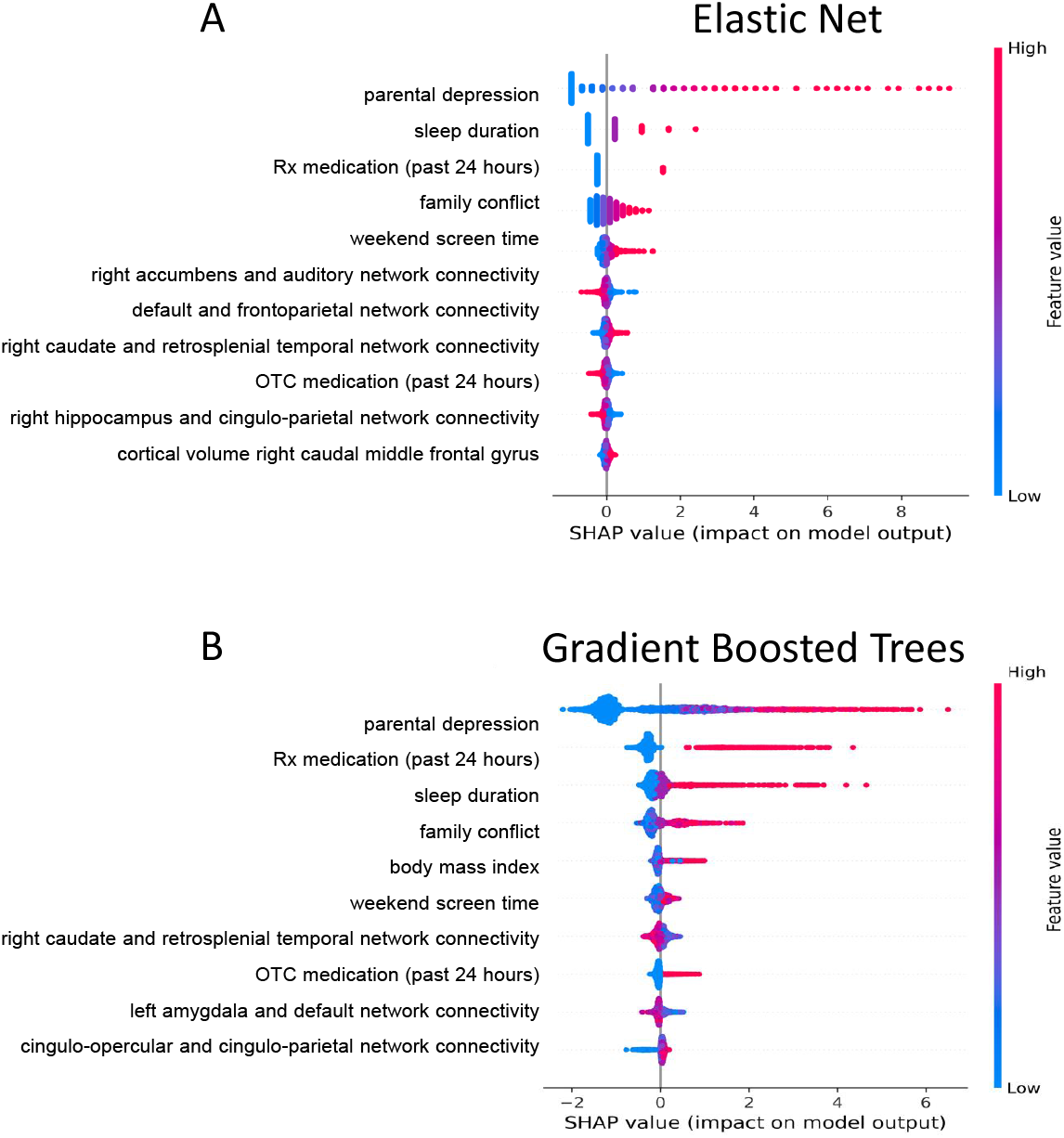
Shapley values of the top 10 features in the elastic net (A) and gradient boosted trees (B) models predicting 1-year follow-up depression symptoms. The summary plots indicate the relationship between the value of a feature and the impact on the prediction, thus combining feature importance with feature effects. Each point on the summary plot is a Shapley value for a feature and an instance. The position on the y-axis is determined by the feature and on the x-axis by the Shapley value. The color represents the value of the feature from low (blue) to high (pink). The features are ordered according to their importance. See **Figure S3** for a summary of the magnitude of Shapley values per feature in each model.

With respect to neural features, resting-state functional connectivity between the right caudate and retrosplenial-temporal network (RTN) was the 5^th^ most important neural feature in the EN model whereas resting-state functional connectivity between the right caudate and visual network was the 6^th^ most important neural feature in the GBT model. With respect to neural features predicting depression symptoms at one-year follow-up, resting-state functional connectivity across a broader set of networks was identified in the EN model (including connectivity between the right caudate and RTN, which was the 3^rd^ most important neural feature), whereas in the GBT model, resting-state functional connectivity between the right caudate and visual network remained the most important neural feature (see **Figures 2-3** and **Figures S2-S3**). Parental depression and family conflict were moderately correlated (*r*=0.29), as was sleep duration and weekend screen time (*r*=0.21); see **Figure S4** for a zero-order correlation matrix among the top features in these models using Pearson’s correlation.

With the exception of cortical volume of the caudal portion of the middle frontal gyrus in the GBT models (see **Figures 2-3**), structural MRI features were not identified among the top contributing features in our predictive models. In a supplemental analysis, we ran an EN model and a GBT model with demographic, clinical, and resting-state fMRI features only to predict baseline and follow-up CBCL scores; these models performed similarly to their full model counterparts (baseline EN: MAE=3.628, *R*^*2*^=0.220; follow-up EN: MAE=4.244; *R*^*2*^=0.101; baseline GBT: MAE=3.666; *R*^*2*^=0.194; follow-up GBT: MAE=4.032; *R*^*2*^=0.122). Moreover, many of the same top features, including parental depression, sleep duration, prescription medication usage in the past 24 hours, weekend screen time, family support, being of mixed race, and resting-state fMRI connectivity between the right caudate and the RTN, were identified in these models. Our results therefore suggest that structural MRI features did not contribute meaningfully to the prediction of CBCL depression scores (either at baseline or follow-up).

## Discussion

This is the first investigation utilizing the ABCD data to examine structural and functional MRI predictors of concurrent and subsequent (one-year follow-up) child depression symptoms. We found that higher levels of parental symptoms of depression, higher levels of family conflict, fewer hours of sleep per night (i.e., sleep duration), and medication usage were among the strongest features contributing to prediction of depression symptoms across two distinct machine learning modeling approaches. While resting-state functional connectivity of several cortical networks were weak contributors in the elastic net model, lower resting-state functional connectivity between the right caudate and retrosplenial-temporal network (RTN) was the most important neural feature (and 5th most important overall feature) in the elastic net (EN) model, while resting-state functional connectivity between the right caudate and visual network was the most important neural feature (and 6^th^ most important overall feature) in the gradient boosted trees (GBT) model. Interestingly, lower connectivity between the right caudate and RTN was the strongest neural feature across both modeling approaches in predicting depression severity in the subset of participants (*n*=7,998) who completed the one-year follow-up assessment. Parental depression and family conflict were moderately correlated (*r*=0.29), as was sleep duration and weekend screen time (*r*=0.21); the remaining top features showed weaker or null correlations, suggesting that the family environment (parental depression and family conflict), sleep quality (sleep duration and weekend screen time), and functional neural factors represent distinct predictors of depression. Together, our results highlight family environment and sleep quality— both of which are modifiable processes—as important risk factors for adolescent depression.

Moreover, our results also demonstrate that lower resting-state functional connectivity of the right caudate with regions comprising visual attention and default mode network functioning— including processing underlying autobiographical memories and self-reflection—may be a promising biomarker of early depression symptoms in adolescence.

Parental mental health and relationship quality between the child and parent remain one of the strongest predictors of mental health outcomes in adolescents, particularly for depression (39). Researchers have identified a multitude of pathways that may explain the intergenerational transmission of depression—including genetic as well as environmental (e.g., caregiving quality, epigenetic) factors—although the relative contribution of these purported mechanisms still remains unknown (for reviews, see references (18, 40, 41). Critically, there is evidence of biological alterations in youth whose parents have a history of depression, including smaller gray matter volumes in subcortical structures (42, 43), HPA-axis dysfunction (for a review see 40), and alterations in epigenetic markers (45, 46), even when youth whose parents have a history of depression but do not endorse current depression symptoms themselves. That higher levels of family conflict were identified as an important predictor of both current and prospective symptoms of depression further supports the formulation that parental behaviors, which are informed by mental health status, are critical for scaffolding offspring brain development. It will be important for future investigations of the ABCD data to examine possible mechanisms (e.g., caregiving style) that could be targeted to mitigate risk for depression.

Our investigation also identified shorter sleep duration and longer weekend screen time usage as important features for predicting depression symptoms. Adolescence is an important time of development that includes changes in chronobiological processes that regulate circadian rhythms, as well as changes in sleep homeostasis that, together, affect the timing and duration of sleep (47). Our findings are consistent with several studies have found that shorter sleep duration is associated with higher depression and other mental health symptoms in adolescents (48, 49).

Indeed, previous analyses from the ABCD dataset using non-regularized linear models found that depressive symptoms were significantly correlated with shorter sleep duration and, furthermore, that greater depression symptoms at baseline predicted shorter sleep duration at follow-up (50). We replicate these findings in a larger sample and further highlight the potential role of screen time. Although there has been controversy regarding the extent to which screen time affects mental health in adolescents (see 42–44), our results indicate that more weekend screen time, which will likely reflect more leisure usage, is related to more depression symptoms at baseline and is predictive of greater depression symptoms one-year later. Together, our results suggest that sleep quality may be targetable processes in reducing depression risk. An important next step in this line of research will be obtain richer contextual information to understand the different sources of shorter sleep duration and longer screen time usage (e.g., academic pressure), and if these sources moderate the associations between sleep quality and depression.

While several resting-state functional connectivity features contributed weakly to the prediction of depression symptoms, lower resting-state functional connectivity between the right caudate and retrosplenial-temporal and visual networks remained the most consistent neural features in predicting depression symptoms. Our findings are broadly consistent with a meta-analytic investigation that identified modest gray matter volume reductions in the caudate (Cohen’s *d*=−0.31) in depressed versus psychiatrically healthy adults (54) and in empirical studies that have identified lower structural connectivity of a caudate-based network in depressed versus psychiatrically healthy adults (55) and adolescents (56). As a component of the dorsal striatum, the caudate plays an important role in reward processing and stimulus-behavior mapping, particularly in contexts where stimulus outcomes are perceived to be contingent on one’s own behavioral actions (57). While previous studies have identified reward-related signals in the ventral striatum as being informative for predicting depression symptoms in community youth (10), our results suggest that task-independent connectivity of the caudate may also be a useful risk marker for depression, with the added advantage of not requiring a potentially demanding cognitive assessment.

In contrast, structural MRI features largely did not contribute to any of the models predicting concurrent or future depression symptoms. Previous investigations of the ABCD data using non-regularized linear models found that smaller cortical surface area (but not cortical thickness) and volumes of regions in the default mode network (ventromedial prefrontal cortex, precuneus, and posterior cingulate cortex) in lateral and medial orbitofrontal cortex (OFC), and in frontal, temporal, and motor regions was associated with greater concurrent depression symptoms (50). Similarly, studies from the ENIGMA Major Depressive Disorder (MDD) consortium (12), which utilizes meta-analytic techniques to estimate the effects of depression on FreeSurfer-derived brain morphometry across different sites worldwide, have also reported similar findings.

Specifically, in one subanalysis that compared 294 healthy controls to 213 depressed adolescents, the investigators found that depressed adolescents exhibited lower surface area (but no differences in cortical thickness), with the strongest effect sizes in the medial OFC, superior frontal gyrus, and visual, somatosensory, and motor areas (12). Consistent with our results, the studies from ENIGMA MDD did not find evidence that subcortical and cortical morphometry was associated with current depression symptoms (12, 13, 15). Importantly, our study extends these previous “big data” investigations that focused on examining associations between regional surface area and adolescent depression by demonstrating that functional connectivity patterns of the right caudate with visual attention and default mode network regions contribute to the prediction of concurrent and future symptoms of depression and may, thus, represent potentially more sensitive biomarkers of early depression symptoms than structural markers.

Several limitations must be considered when interpreting our findings. First, while the CBCL provides dimensional measures of depression symptoms, it is a measure based on parent-report, which may not necessarily be concordant with child-report, particularly for depression and other internalizing disorders (58, 59). Future assessments of the ABCD Study will also include the Youth Self-Report, a child-reported questionnaire that captures the same symptom dimensions as the CBCL, which will provide us an opportunity to replicate the analyses in the present investigation. Similarly, sleep duration and screen time were measured based on parent-report from questionnaires; future assessments of the ABCD Study will seek to comprehensively assess these constructs using more objective measures (e.g., actigraphs or other wearables, sensor data from smartphones) in order to understand the role of sleep quality on depression risk in adolescents.

In conclusion, our study identified multi-level predictors of depression symptoms in a nationally representative sample of over 8,500 youth ages 9-10. We found that higher parental depression symptoms, higher levels of family conflict, shorter sleep duration, longer weekend screen time, and lower resting-state functional connectivity of the caudate was associated with greater concurrent and one-year depression symptoms. Our findings point to two modifiable processes for depression risk that may be important considerations for depression prevention—family environment and sleep quality—as well as highlight the potential of caudate-based functional connectivity patterns to be biomarkers of early depression emergence in youth.

## Data Availability

Data used in the preparation of this article were obtained from the Adolescent Brain Cognitive Development (ABCD) Study (https://abcdstudy.org), held in the NIMH Data Archive. The ABCD consortium investigators designed and implemented the study and/or provided data but did not necessarily participate in the analysis or writing of this report. This article reflects the views of the authors and may not reflect the opinions or views of NIH or of the ABCD consortium investigators. The ABCD data repository grows and changes over time. The ABCD data used in this report came from https://dx.doi.org/10.15154/1504431

## Acknowledgments

The ABCD Study is funded by NIH and other federal partners under grants DA-041048, DA-050989, DA-051016, DA-041022, DA-051018, DA-051037, DA-050987, DA-041174, DA-041106, DA-041117, DA-041028, DA-041134, DA-050988, DA-051039, DA-041156, DA-041025, DA-041120, DA-051038, DA-041148, DA-041093, DA-041089, DA-041123, and DA-041147. A full list of supporters is available at https://abcdstudy.org/federal-partners.html.

This research was supported by the National Institutes of Health (K01MH117442 to TCH), the Ray and Dagmar Dolby Family Fund (to TCH), and the National Institute on Alcohol Abuse and Alcoholism (F31AA027169 to ACM). The funding agencies played no role in the design and conduct of the study; collection, management, analysis, and interpretation of the data; and preparation, review, or approval of the manuscript. All authors report no biomedical conflicts of interest.

