## Supplement for "Multi-level Predictors of Depression Symptoms in the Adolescent Brain Cognitive Development (ABCD) Study"

*Image Processing*

Brain features derived from structural MRI included subcortical gray matter volumes, cortical thickness, and cortical surface area from regions defined by the Desikan atlas (Desikan et al., 2006) using *recon-all* from FreeSurfer version v. 5.3.0 (<http://surfer.nmr.mgh.harvard.edu/>; Fischl et al., 2002). As described in more detail in Hagler et al. (2019), this processing pipeline included removal of non-brain tissue, segmentation of subcortical white matter and gray matter structures used an automated atlas-based procedure, and cortical parcellations, where cortical gray matter and underlying white matter voxels were labeled according to surface-based nonlinear registration to the atlas based on cortical folding patterns (Fischl et al., 1999b) and Bayesian classification rules (Desikan et al., 2006; Fischl et al., 2004). Structural MR images that did not pass quality control procedures (including visual inspection of raw images for artifacts or poor image quality, visual inspection of the FreeSurfer cortical reconstruction and segmentation results, a neuroradiological read for incidental findings, and other quality control metrics such as signal-to-noise ratio, head motion) by the ABCD team were excluded (see Table S1).

Brain features derived from resting-state structural fMRI included functional connectivity between the networks defined by the Gordon parcellation (Gordon et al., 2014) and also between each of the subcortical structures segmented by FreeSurfer (see above) and the Gordon networks using seed-based functional connectivity adapted for cortical surface-based analyses (Seibert and Brewer, 2011). Specifically, regions within the Gordon parcellation were classified as belonging to a particular network or community (e.g., restroplenial temporal, salience network); thus, average correlations within a network were computed as the average of correlations (after Fisher-r-to-z transformation) for each pairwise combination of regions with membership to said network. Similarly, correlations between each network and each subcortical structure segmented by FreeSurfer by averaging the correlations (after Fisher-r-to-z transformation) between each of these regions. As described in more detail in Hagler et al. (2019), preprocessing of the resting-state fMRI data included correcting for B_0_ distortions, estimating framewise displacement (FD) and temporal SNR. Participants for whom more than 10% of their timeseries had FD > 0.2 mm or for whom there was fewer than 375 usable timepoints for modeling were excluded from the present analyses.

*Features (Predictors)*

Sleep duration was measured from a single item from the Sleep Disturbances Scale for Children (see **Table S1** for the filename containing responses from this measure), which asks parents, “How many hours of sleep does your child get on most nights?” (for the past 6 months). Responses on this item range from 1-5, where 1 indicates that the child sleeps 9-11 hours per night, 2 indicates that the child sleeps 8-9 hours per night, 3 indicates that the child sleeps 7-8 hours per night, 4 indicates that the child sleeps 5-7 hours per night, and 5 indicates that the child sleeps less than 5 hours per night. The value range for this question is 1, 2, 3, 4, 5, where 1 indicates that a child sleeps 9-11 hours per night; 2 indicates that a child sleeps 8-9 hours per night; 3 indicates that a child sleeps 7-8 hours per night; 4 indicates that a child sleeps 5-7 hours per night; and 5 indicates that a child sleeps less than 5 hours per night. Thus, a higher score on this measurement indicates a shorter amount of sleep.

Family conflict was operationalized based on the sum of the first 9 items from the Family Environment Questionnaire, as these were the items used in all timepoints of the ABCD study. Higher scores on this scale indicate more family conflict. A summary of these items can be found in **Table S2**.

*Missing Data*
All subjects were missing structural MRI data for the following variables: smri_vol_scs_lesionlh, smri_vol_scs_lesionrh, smri_vol_scs_wmhintlh, smri_vol_scs_wmhintrh. These variables were therefore not included in our analyses as features. Out of the total 11,878 subjects, 142 subjects were missing structural MRI data and 571 subjects were missing resting-state fMRI data and were therefore excluded from our analyses. Additional subjects were excluded based on poor quality control metrics and/or excessive motion in the structural and fMRI data (see “Image Processing” above for more details). For all twins or triplets, we randomly selected one subject within each family unit to be included in our analyses. Finally, subjects without CBCL scores at baseline were also excluded from all analyses. Please see **Figure S1** for a comprehensive flowchart from the full dataset to the final analytic sample.

***Feature Importance***

We used the Shapely Additive exPlanations (SHAP) method to compute feature importance scores for each feature in our models. A Shapley value is the average marginal contribution of a feature value across all possible combinations of features without the given feature. Let us consider a dataset with **n** features. The ***i^th^*** feature’s Shapely value will be the mean marginal contribution over all possible combinations of the remaining **n-1** features. As the total number of ways of selecting any number of objects out of a set of **r** objects is **2^r^**, **n*****2^n^** values are computed to determine the marginal contribution of every feature (**2^n^** per feature x ***n*** features). Because the computation of Shapley values rises exponentially with the number of features, our total feature set (398 features) rendered us with unviginticentillion (~10^122^) calculations, which is impossible to complete even on a supercomputer. Thus, we employed the SHAP as a computationally efficient want to compute Shapely values. While the details of its algorithmic implantation is beyond the scope of this paper, briefly, we used a linear explainer to approximate Shapely values for the EN models (a method which accounts for multi-collinearity among features) and a tree explainer to approximate Shapely values for the GBT models (a method which uses ensembles of trees under several possible feature dependence assumptions).

**Figure S1. Flowchart of study participants included in final analytic sample (N=8,507).**

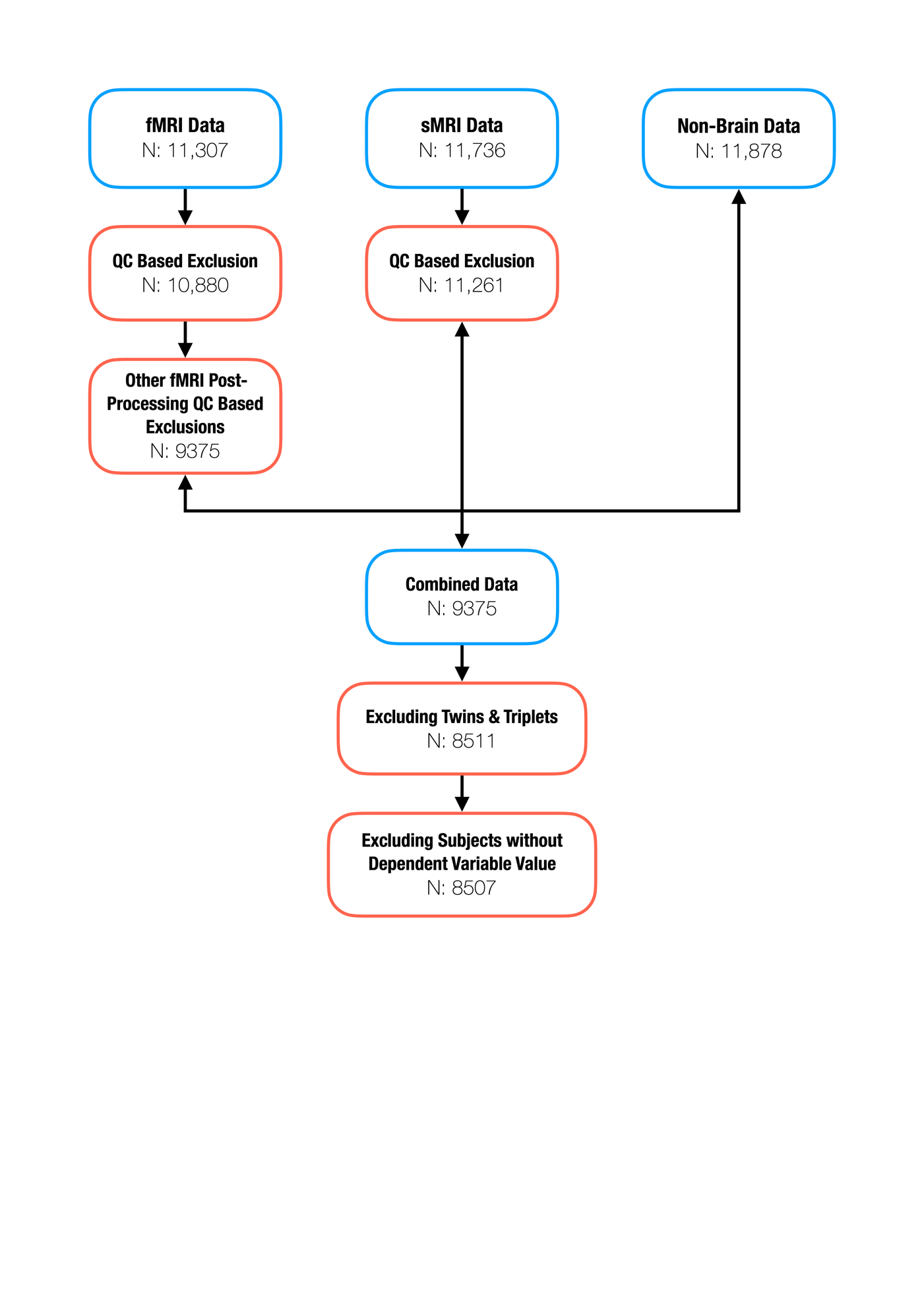

**Figure S2. Magnitude of feature importance values for top 10 features in the elastic net (A) and gradient boosted trees (B) models predicting baseline depression symptoms.** Features sorted according to importance (absolute Shapley value). Note that after feature #4, there is a sharp drop in the impact of the subsequent features in contributing to model performance.

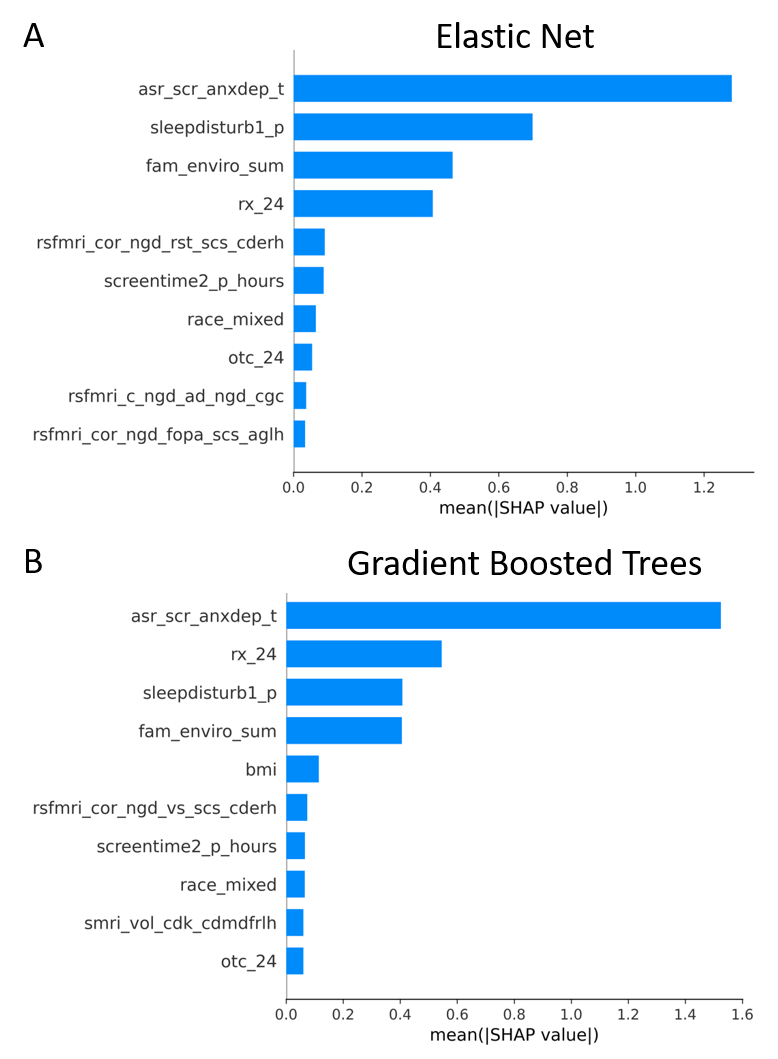

**Figure S3. Magnitude of feature importance values for top 10 features in the elastic net (A) and gradient boosted trees (B) models predicting 1-year depression symptoms.** Features sorted according to importance (absolute Shapley value). Note that after feature #5, there is a sharp drop in the impact of the subsequent features in contributing to model performance.

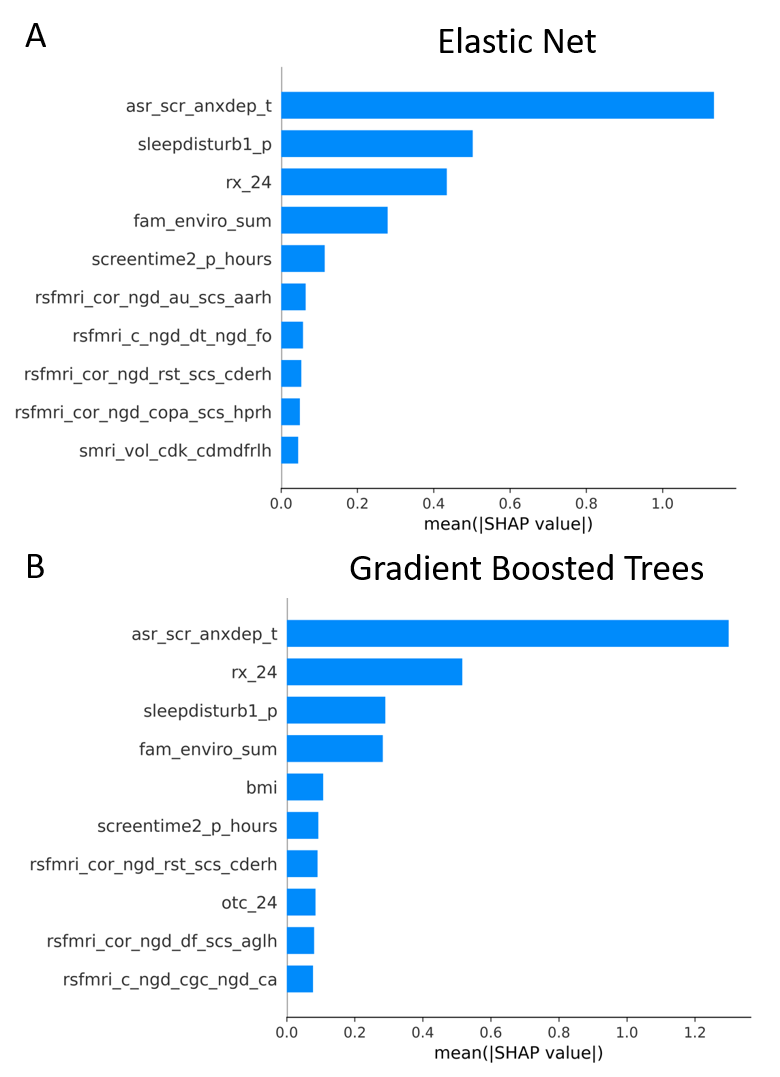

**Figure S4. Correlation values among the most predictive features in our models.** fam_enviro_sum=family conflict; sleepdisturb1_p=weekend screen time; rsfmri

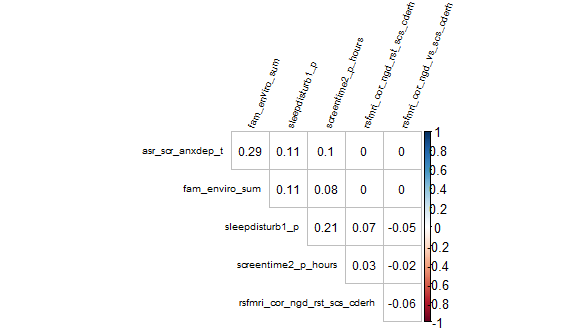

**Table S1. Comparison of demographic and related variables between the full sample (N=8,507) and the sample with follow-up data (N=7,998).** All variables provided here refer to the values at baseline.

|  | **Full sample** | **Follow-up sample** |
| --- | --- | --- |
| **N** | 8507 | 7998 |
| **Sex (% Female)** | 49.65% | 49.52% |
| **Age (years)** | 9.49 ± 0.51 (8 – 11) | 9.49 ± 0.51 (8 – 11) |
| **Race (% White)** | 75.23% | 76.36% |
| **Puberty Development Score** | 6.77 ± 2.89 (1 – 20) | 6.74 ± 2.87 (1 – 20) |
| **BMI** | 18.98 ± 32.75 (2.08 - 52.82) | 18.96 ± 33.76 (2.08 - 52.82) |
| **Caffeine Consumption in the past 24 hours (% yes)** | 17.85% | 17.44% |
| **Prescribed Medications taken in the past 24 hours (% yes)** | 14.04% | 14.25% |
| **Over-the-counter Medications taken in the past 24 hours (% yes)** | 14.85% | 15.25% |
| **Sleep Duration**  **(Number of Hours of Sleep Per Night)** | 9-11 hours: 4072 (47.87%)  8-9 hours: 3181 (37.39%)  7-8 hours: 969 (11.39%)  5-7 hours: 263 (3.09%)  <5 hours: 22 (0.26%) | 9-11 hours: 3897 (48.72%)  8-9 hours: 2976 (37.21%)  7-8 hours: 870 (10.88%)  5-7 hours: 236 (2.95%)  <5 hours: 19 (0.24%) |
| **Child Education (% Endorsed)** | 1^st^ grade: 0.025%  2^nd^ grade: 0.45%  3^rd^ grade: 17.03%  4^th^ grade: 44.62%  5^th^ grade: 34.71%  6^th^ grade: 3.14%  7^th^ grade: 0.025% | 1^st^ grade: 0.03%  2^nd^ grade: 0.39%  3^rd^ grade: 17.12%  4^th^ grade: 44.61%  5^th^ grade: 34.75%  6^th^ grade: 3.08%  7^th^ grade: 0.02% |
| **Parental Marital Status (% Endorsed)** | Married: 69.39%  Never Married: 11.66%  Divorced: 8.73%  Living with Partner: 5.65%  Separated: 3.79%  Widowed: 0.78%  [60] | Married: 70.59%  Never Married: 10.97%  Divorced: 8.62%  Living with Partner: 5.45%  Separated: 3.61%  Widowed: 0.76% |
| **Total Combined Family Income for the past 12 months (% Endorsed)** | <$5k: 3.08%  $5k - $12k: 3.25%  $12k - $16k: 2.45%  $16k - $25k: 4.28%  $25k - $35k: 5.70%  $35k - $50k: 7.81%  $50k - $75k: 12.61%  $75k - $100k: 22%  $100k - $200k: 28.06%  >$200k: 10.76% | <$5k: 2.81%  $5k - $12k: 2.99%  $12k - $16k: 2.30%  $16k - $25k: 4.06%  $25k - $35k: 5.50%  $35k - $50k: 7.65%  $50k - $75k: 12.68%  $75k - $100k: 21.96%  $100k - $200k: 28.87%  >$200k: 11.18% |
| **Highest Parental Education (% Endorsed)** | 3^rd^ grade: 0.03%  4^th^ grade: 0.02%  5^th^ grade: 0.02%  6^th^ grade: 0.38%  7^th^ grade: 0.11%  8^th^ grade: 0.34%  9^th^ grade: 0.73%  10^th^ grade: 0.71%  11^th^ grade: 0.99%  12^th^ grade (no diploma): 1.39%  High school graduate: 6.62%  GED or equivalent: 2.62%  Some College Degree: 12.80% Associate Degree (Occupational, Technical, or Vocational): 7.41%  Associate Degree (Academic Program): 5.2%  Bachelor’s Degree: 25.30%  Master’s Degree: 24.40% Professional School Degree: 5.17%  Doctoral Degree: 5.76% | 3^rd^ grade: 0.04%  4^th^ grade: 0.02%  5^th^ grade: 0.03%  6^th^ grade: 0.34%  7^th^ grade: 0.11%  8^th^ grade: 0.34%  9^th^ grade: 0.70%  10^th^ grade: 0.69%  11^th^ grade: 0.83%  12^th^ grade (no diploma): 1.29%  High school graduate: 6.25%  GED or equivalent: 2.36%  Some College Degree: 12.29% Associate Degree (Occupational, Technical, or Vocational): 7.30%  Associate Degree (Academic Program): 5.11%  Bachelor’s Degree: 25.82%  Master’s Degree: 25.12% Professional School Degree: 5.34%  Doctoral Degree: 6.02% |
| **Family Environment Score** | 2.52 ± 1.96 (0 – 9) | 2.51 ± 1.96 (0 – 9) |
| **Weekend daily screen time (hours)** | 3.80 ± 2.61 (0 – 24) | 3.78 ± 2.58 (0 – 24) |
| **CBCL Depression (t-score)** | 53.56 ± 5.69 (50 – 89) | 53.87 ± 6.02 (50 – 87) |
| **Adult Self-Report Depression (t-score)** | 53.43 ± 5.70 (50 – 98) | 53.40 ± 5.63 (50 – 96) |

**Table S2. Summary of ABCD file names where features and outcome variables were extracted.**

| **Variables** | **Data Structure File Name(s)** |
| --- | --- |
| BMI | abcd_ant01 |
| CBCL | abcd_cbcls01 |
| Demographics (e.g., age, education) | pdem02 |
| Familial Relation | acspsw03 |
| Family Environment Scale | fenvs01 |
| FreeSurfer Quality Control | freesqc01 |
| rsfMRI Quality Control | abcd_mrirstv02 |
| Functional MRI | abcd_betnet01 (cortical-cortical), mrirscor02 (subcortical-cortical) |
| MRI Info (e.g., scanner type) | abcd_mri01 |
| Pubertal Stage | abcd_ppdms01 |
| Screen time | stq01 |
| Site | abcd_lt01 |
| Sleep | abcd_sds01 |
| Structural MRI | abcd_smrip101 (cortical), abcd_smrip201 (subcortical) |
| Substances taken in the past 24 hours | medsy01 |

**Table S3. Items from the Family Environment Scale.** All items are rated as “True” or “False” and coded as “0” and “1”, respectively. Items denoted with * are reversed scored. Higher scores on this measure indicate more family conflict.

**Item**

Family members really help and support one another

We fight a lot in our family*

Family members rarely become openly angry

We often seem to be killing time at home*

We put a lot of energy into what we do at home

There is a feeling of togetherness in our family

Family members sometimes get so angry they throw things*

Family members rarely ever lose their temper

We rarely volunteer when something has to be done*
